# Access to social protection by people living with, at risk of, or affected by HIV in Eswatini, Malawi, Tanzania and Zambia - results from Population-Based HIV Impact Assessments

**DOI:** 10.1101/2021.10.11.21264851

**Authors:** David Chipanta, Audrey Pettifor, Jessie Edwards, Danielle Giovenco, Hillary Mariko Topazian, Rachel M. Bray, Monique C. Millington, Janne Estill, Olivia Keiser, Jessica E Justman

## Abstract

We aimed to measure social protection (SP) coverage among the general population and women and men living with HIV (WLHIV, MLHV), female and male sex workers (FSW, MSW), men who have sex with men (MSM), adolescent girls young women (AGYW), and orphans vulnerable children (OVC). We used Population-Based HIV Impact Assessments data from Eswatini, Malawi, Tanzania and Zambia. We generated survey-weighted proportions for each population group receiving any SP benefits, along with 95% confidence intervals (CI) using jackknife variance estimation. The proportion reported receiving SP benefits among the general population ranged from 7.7% (95% CI: 6.7%–8.8%) in Zambia to 39.6% (95% CI: 36.8%– 42.5%) in Eswatini. SP benefits by WLHIV, MLHIV, AGYW, OVC, SW and MSM – were lower than the 2017-19 global average of 45%. Data on access to SP benefits by people living with or affected by HIV from other regions is needed to estimate their SP coverage better.

## INTRODUCTION

In 2016, the United Nations Member States adopted the Political Declaration on Ending AIDS by 2030. The declaration laid out commitments to reduce new HIV infections to fewer than 500,000, AIDS-related deaths to less than 500,000 and eliminate HIV-related stigma and discrimination globally [1]. Summarized in the Ten UNAIDS Fast-Track Commitments, if achieved by 2020, the world would be on course to ending AIDS as a public health threat by 2030 [1,2]. With the COVID-19 pandemic risking setting back progress against HIV, effectively implementing and measuring these commitments is crucial to the AIDS response [3,4]. Most of the Ten UNAIDS Fast-Track Commitments have been measured. However, Commitment 6, “Ensure that by 2020, 75% of people living with, at risk of or affected by HIV benefit from HIV-sensitive social protection,” has not. Social protection has recently achieved increased attention because it is integral to the COVID-19 pandemic response. More than 3,330 new social protection programmes worth US$2.9 trillion have been introduced globally since 2020 to mitigate the health, social and economic fallout from the COVID-19 pandemic [5]. More than cash transfers, social protection comprises private and public transfers, and policies, such as social safety nets, social security and labour market policies to help people manage risk and protect them from poverty and destitution [6,7]. People living with HIV (PLHIV), key populations (sex workers [SW], gay men and other men who have sex with men [MSM], people who inject drugs, transgender people and prisoners), adolescent girls and young women (AGYW), orphans and vulnerable children (OVC) and others have increasingly demanded access to social protection benefits [8–10].

Social protection impacts can be powerful, especially in settings where people face multiple threats to their health and well-being [11-19]. It impacts HIV prevention and treatment outcomes through complex pathways. Social protection enables people to withstand life shocks, empowering them to adopt less HIV risky coping strategies [14,15,19,20]. Regarding affordability and access to ART and health care, social protection helps people overcome financial and other barriers to HIV treatment and prevention services, reduces inequity in accessing and using services [12,15,20]. By helping enrol and keep adolescent girls and young women in school, social protection protects them from HIV [11,14,15,17–20]. It is also critical for carers, many of whom are women, in unpaid caregiving work [20]. Social protection policies and laws, including workplace policies, uphold the rights to gainful employment, social security, housing, non-discrimination and other social and economic rights of everyone [21,22].

Several features of social protection implementation in sub-Saharan Africa are crucial to HIV prevention and treatment efforts. First, countries, including those heavily affected by HIV, are scaling up social protection programmes to help people overcome shocks such as climate change, price increases, and most recently COVID-19 pandemic [5,23]. Shocks interrupt HIV services [4]. Pandemic related social protection help mitigate the vulnerability of people living with, at risk of or affected by HIV. Second, cash transfers are a primary delivery mechanism of social protection [7,24]. Because of their multiple impacts on health and development domains, cash transfers have heightened attention in HIV prevention and treatment efforts [15,16,18,25]. Third, children, adolescents, girls and women, people with disabilities, and older people, are often the focus populations of social protection programmes [7,24]. These are also affected by HIV. Fourth, social protection, including floors, is promoted as a human right, applicable to everyone, including people living with, at risk of or affected by HIV [21,22]. Thus, measuring social protection coverage is crucial for the AIDS response.

In 2017 the UNAIDS National Composite and Policy Index (NCPI) first reported data on social protection [26]. This NCPI social protection data reflect whether social protection strategies are HIV sensitive, that is, whether they refer to HIV or recognize PLHIV, key populations, AGYW, OVC and people affected by HIV as crucial beneficiaries; and whether they address unpaid work in the HIV context [19,20]. However, none of these elements measure Commitment 6 directly. This study aimed to measure Commitment 6 by estimating social protection coverage among the general population and seven sub-population groups who are living with, at risk of or affected by HIV: women and men living with HIV (WLHIV, MLHIV), female and male sex workers (FSW, MSW), MSM, AGYW and OVC. We used publicly available Population-Based HIV Impact Assessment (PHIA) data from four high HIV prevalence countries: Eswatini, Malawi, Tanzania and Zambia.

## METHODS

### Study setting

Table 1 displays selected economic indicators, HIV estimates and the proportion of people who reported receiving social protection benefits in the four study countries. The countries are located in Eastern and Southern Africa, the epicentre of the HIV epidemic. Eswatini, Tanzania and Zambia are lower middle-income countries. Malawi is a low-income country. Eswatini and Zambia are among the countries with the highest HIV prevalence and incidence worldwide. Of the four countries, they are also the most unequal in terms of wealth. Their Gini coefficients, a measure of inequality, were 54.6 and 57.1, respectively, on a scale of zero (total equality) to 100 per cent (full inequality) [27]. All four countries have established national social protection programmes and mature HIV epidemics [6,28].

**Table 1.**
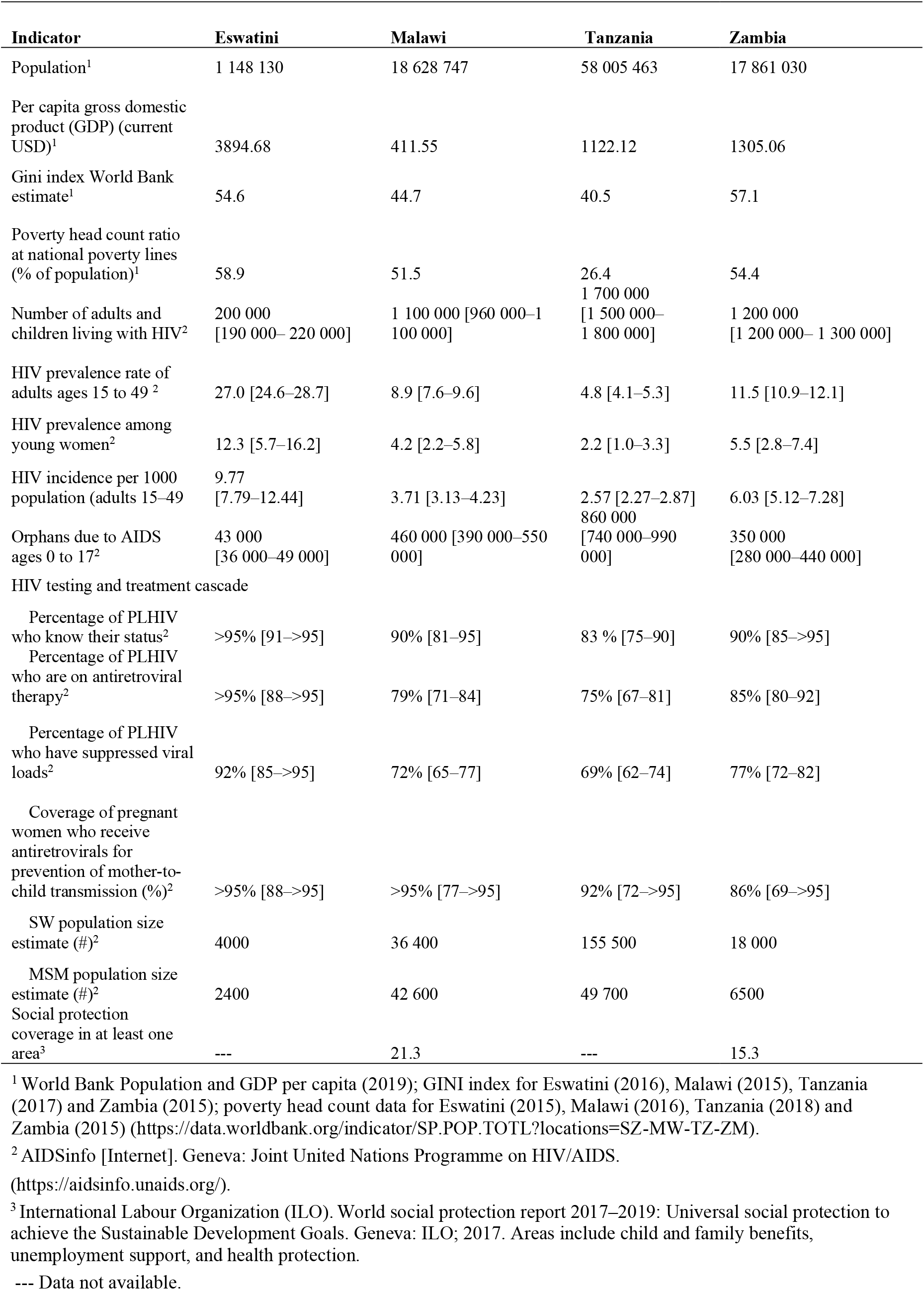
Per capita gross domestic product, Gini index, poverty ratio, HIV estimates and proportion of people who reported accessing any social protection benefit, by country.

As of 2020, Eswatini had fewer PLHIV (200,000) than Malawi (1,100,000), Tanzania (1,700,000) or Zambia (1,200,000) [28]. Tanzania had the largest estimated number of SW (155,500) and MSM (49,700), followed by Malawi (36,400 SW; 42,600 MSM), Zambia (18,000 SW; 6500 MSM) and Eswatini (4000 SW; 2400 MSM) [28].

### Data sources

We used Indicator 1.3.1, Proportion of population covered by at least one social protection benefit, of the Sustainable Development Goals (SDGs) [29], which has been identified by UNAIDS as a proxy to measure Commitment 6.

We analysed data from the PHIA surveys that were publicly available and had data on social protection coverage among OVC, AGYW, PLHIV, SW and MSM. The PHIA surveys measured the impact of HIV programs in countries supported by the United States President’s Emergency Plan for AIDS Relief. These cross-sectional surveys were administered to consenting individuals in nationally representative random cluster samples of households. Study procedures included administering questionnaires, household-based HIV counselling and testing and immediate return of point of care test results. The surveys assessed HIV status and included questions about external economic support and engagement in sex work for men and women. They identified AGYW ages 15 to 24 years directly, and indirectly, MSM and OVC status through behavioural and family demographic questions. Data on HIV and social protection variables were completed and available for four surveys, allowing for standardized measures across the corresponding countries [30]: Eswatini (2016–2017), Malawi (2015–2016), Tanzania (2016–2017) and Zambia (2016). We obtained the PHIA data sets from the PHIA Project website at https://phia-data.icap.columbia.edu/files.

We used the Household, Adult and Child Interview and Adult HIV Biomarker data sets. In participating households, a household questionnaire was administered to the household head, who indicated all individuals living in the household (referred to as the roster or household list). Then, individual questionnaires were administered to eligible and consenting individuals in the household. Adults (15 years and older) completed an adult questionnaire. Adults also provided data on their children ages 0–14 years as part of the “children” module of the adult questionnaire. The Adult HIV Biomarker data set contained HIV test results of all adults and adolescents aged 15 and older who completed an individual interview and consented or assented to provide blood samples for HIV testing. The child interview data set included variables from the roster, such as age and gender, and questions from the adult questionnaire’s children module that were attached to the child’s records [30].

### Variables and outcome descriptions

We included women and men (15 to 59 years old) who were interviewed. We defined the respondent as HIV positive if their HIV biomarker test was positive. SW were defined as males or females aged 15 years or more who reported selling sex for money in the past 12 months; and AGYW were defined as females 15–24 years of age. We defined a person as MSM if the respondent was male and their first, second or third most recent sexual partner in the last 12 months was male. We also included OVC, defined as children ages 0–17 years who were orphaned, or HIV-positive, lived in a household with chronically ill parents or experienced a recent death from chronic illness. If gender was missing, the person was excluded from the analysis. Analyses of adults included only those adults interviewed, whereas all children in the roster ages 0–17 years were eligible for inclusion if the household head indicated that they were OVC.

We considered any external economic support to the household in the last three or 12 months. We recorded the receipt of any child support provisions if the respondent acknowledged receiving any support, including school, social, material, emotional or medical support. (See the Supplementary Table 1 in Appendix 1 for details on how the variables were coded.)

### Analysis

We estimated the proportion receiving any social protection benefits by using the SAS survey means procedure to determine the weighted proportion of persons who reported receiving any social protection benefit for the identified population groups. Survey weights accounting for non-response using Chi-squared automatic interaction detector (CHAID) analysis, non-coverage and the probability of selection were applied. We used individual interview weights in the analyses of adults and household weights in OVC analyses. Variances and 95% confidence intervals (CI) were estimated using the corresponding jackknife replicate weights [31].

We used SAS v9.4 for the analyses [32]. The code is given in the Appendix 2.

## RESULTS

Table 2 shows the sample distribution by country and population groups. The sample percentages are unweighted. We did not aggregate results within and across countries due to the different sampling weights used. The sample comprised 10,233 adults ages 15–59 years in Eswatini, 19,106 in Malawi, 29,638 in Tanzania and 21,278 in Zambia, along with 2,573 OVC ages 0–17 years in Eswatini, 4,471 in Malawi, 7,388 in Tanzania and 6,094 in Zambia. The AGYW population groups comprised between 19% and 22% of the adult sample in each country. OVC accounted for between 19.1% (Malawi) and 27.8% (Eswatini) of children ages 0– 17. MSM and SW accounted for less than one per cent of the sample in each country, except in Tanzania, where FSW made up 2.7%. We did not report estimates for MSM and SW for Eswatini because there were fewer than 25 observations.

**Table 2.**
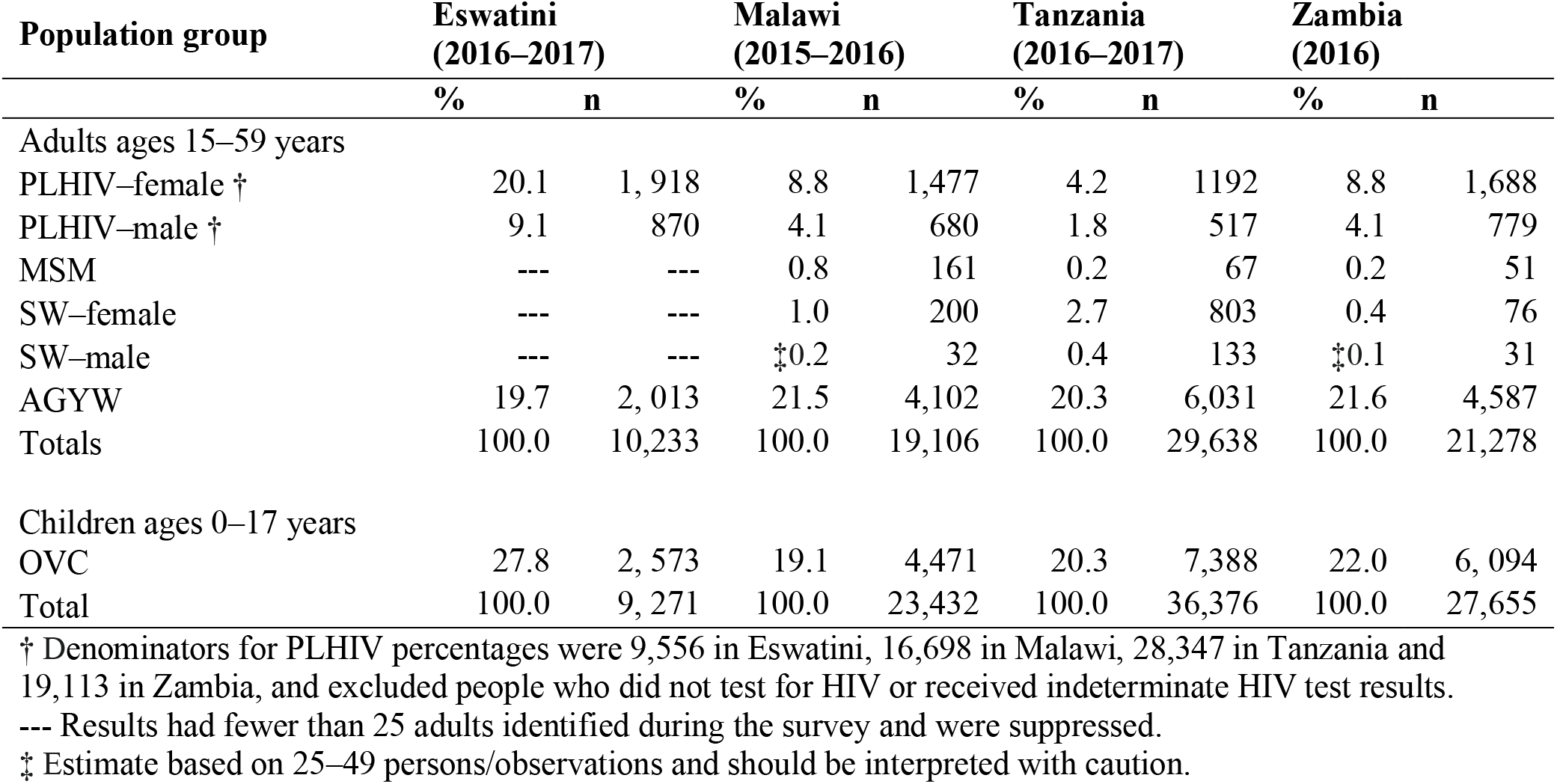
Sample distribution by country and population group, unweighted percentage (%) and size, PHIA.

Among the general population, the proportion who reported receiving any social protection benefits ranged from 7.7% (95% CI: 6.7%–8.8%) in Zambia to 39.6% (95% CI: 36.8%–42.5%) in Eswatini (Table 3). OVC, AGYW, SW, MSM and PLHIV received social protection benefits at a similar level to the general population, with a few exceptions. In Tanzania, 8.8% (95% CI: 7.9%–9.7%) of the general population received social protection benefits, compared with 13.6% (95% CI: 10.8%–16.5%) of WLHIV and 6.1% (95% CI: 4.9%–7.3%) of OVC. However, in Zambia, more OVC (14.4% [95% CI: 12.3%–16.4%] received social protection than the general population (7.7% [95% CI: 6.7%–8.8%]).

**Table 3.**
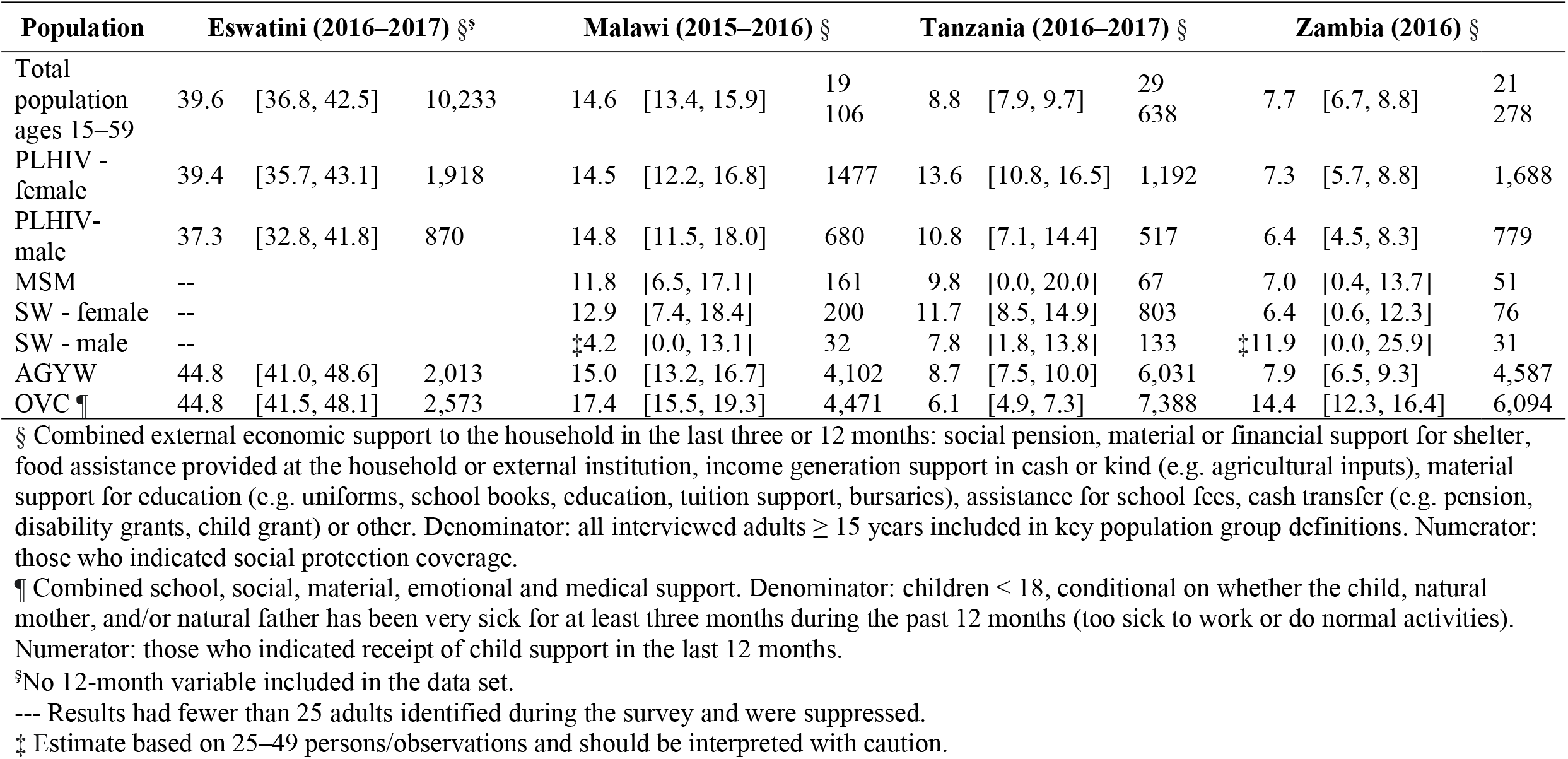
Proportion reporting any external economic support, last 12 months by country and population group, PHIA (%, 95% CI, sample size)

## DISCUSSION

This study found wide variation in the proportion reporting receiving social protection benefits by population group. The proportion reporting receiving social protection benefits was lower than the 2017-19 global average of 45% [6] in all population groups, except for OVC and AGYW in Eswatini. Commitment 6 may have been too ambitious. In general, OVC, AGYW, SW, MSM and PLHIV reported receiving social protection benefits equal to or greater than the general population. In Tanzania, WLHIV reported receiving more social protection benefits than the general population and OVC reported receiving less. OVC reported receiving more social protection benefits in Zambia.

The finding that the proportion of the general population reporting receiving any social protection benefit was lower than the global average and varying widely, from 7.7% (95% CI 6.7–8.8) in Zambia to 39.6% (95% CI: 36.8–42.5) in Eswatini, is consistent with existing evidence. The International Labour Organization (ILO) estimated that only 45 per cent of the global population accessed at least one social protection benefit as of 2019, whereas only 17.8% of Africans reported receiving social protection benefits. About one fifth (21.3%) of the total population in Malawi, 15.3% in Zambia, and 86% and 3.2% of older people (persons above statutory retirement age) in Eswatini and Tanzania, respectively, accessed social protection services based on ILO data [6].

Commitment 6 may have been too ambitious. For example, none of the countries in our study reported any population group accessing social protection benefits more than the 2017-2019 global average of 45%, let alone 75% stipulated in Commitment 6. However, the 2016 Political Declaration from which the Ten UNAIDS Fast-Track commitments are derived stated: “… 75 per cent of people living with, at risk of and affected by HIV *who are in need* [Italics added for emphasis] benefit from HIV-sensitive social protection… “ [1] It focused on a subset of people living with at risk of or affected by HIV in “need” of social protection benefits; not all people living with at risk of or affected by HIV reflected in Commitment 6 [2] and measured by this study. Thus, using the wording of the Political Declaration that includes those in need of social protection benefits, the social protection target might have been achievable. Social protection coverage would have been assessed only among a smaller group of people living with at risk of or affected by HIV in need of social protection benefits; not all of them. Such sub-analysis may still have to be conducted by governments to situate the results in their contexts and identify areas for policy actions.

However, we found that the proportions of OVC, PLHIV (especially women) and AGYW who reported receiving social protection benefits were higher than for SW and MSM. One possible explanation is that community-based organisations and governments in sub-Saharan Africa have developed social protection programmes to mitigate the impact of the HIV epidemic. Children, girls and women have been disproportionately impacted by HIV and are prioritised as populations to reach by many social protection programmes [33]. Zambia’s social protection programmes have historically focused on OVC; Malawi and Tanzania have focused on the poorest households [33]. In addition to these population groups, the focus of Eswatini’s social safety nets has included PLHIV, tuberculosis patients, school-going children and adolescents, and World War II survivors or their dependents [34]. Another consideration is that in the high HIV prevalence countries of Eastern and Southern Africa, OVC, AGYW and PLHIV may be more accepted and considered deserving of socioeconomic support than SW and MSM.

SW and MSM may face stigma and discrimination related to their social identity. They are also criminalised in many countries, preventing them from identifying as SW or MSM. They might avoid accessing services that could lead to disclosing their social identities [9,29,30]. At the same time, SW may be very poor and yet not eligible for government-provided social protection or economic support to small businesses [35,36]. Sex work is not considered as work or as a business in many countries of east and southern Africa. However, in Tanzania, a higher proportion of FSW reported receiving social protection benefits than the general population. They might have been over-represented among women receiving social protection benefits in Tanzania. FSW in Tanzania might have successfully organized themselves to access and provide social protection benefits to each other [37].

The third result from our study was that a larger proportion of PLWHIV, AGYW and OVC groups in Eswatini reported receiving social protection benefits than in Malawi, Tanzania and Zambia. This result is backed by evidence and suggests that a country’s income level plays an essential role in more people receiving social protection benefits [6]. A prosperous country is more likely to provide social protection benefits, including to PLHIV. Eswatini’s per capita GDP is three times that of Tanzania and Zambia and nine times that of Malawi. Spending 1.31 per cent of its GDP, Eswatini fully funded its social assistance programmes; Malawi did not. Malawi spent only 0.41 per cent of its GDP on social assistance programmes [6]. More of Malawi’s people may have depended on limited social assistance, typical among developing countries. Thus, a relatively higher proportion of Malawians reported receiving social protection benefits than the country’s income would suggest [6].

The size of the HIV epidemic and the effectiveness of the HIV response might be crucial in linking people to social protection benefits. Eswatini outperforms Malawi, Tanzania and Zambia on the HIV testing and treatment cascade and has fewer estimated PLHIV. Eswatini’s impressive AIDS response is credited, in part, to an effective multi-sectoral AIDS response coordinated from the Prime Minister’s office by the National Emergency Response Council on HIV/AIDS (NERCHA). NERCHA also directly delivers social protection benefits, including school feeding, food distribution and social services. NERCHA is involved in decision-making about OVC educational grants, supplementary feeding, fee-waivers, agriculture input subsidies, and old age grants delivered by ministries of education, health, agriculture and others. Moreover, Eswatini’s social protection strategies directly include people living with, at risk of and affected by HIV as primary beneficiaries [34]. It has integrated HIV and social protection services within the government. As a result, Eswatini may have done a better job linking people living with, at risk of, or affected by HIV to social protection benefits than Malawi, Tanzania and Zambia. However, people whose social identity is criminalised, such as SW and MSM, may lose out on the benefits, even in relatively richer countries. Focused efforts may be required to enhance access to social protection benefits of people who are criminalised.

To our knowledge, our study is the first to estimate social protection coverage among PLHIV, SW and MSM. We used nationally representative data sets from four countries, enabling us to compare the proportions of seven sub-populations and the general population which reported receiving social protection benefits in four high HIV prevalence countries. United Nations Children’s Fund (UNICEF) developed and piloted social protection questions for indicator SDG in Kenya (2014), Zimbabwe (2015), Vietnam (2015) and Belize (2015), and showed that the questions worked well. UNICEF assessed the adequacy, clarity and relevance of the questions for various population groups and settings [38]. UNICEF did not estimate social protection coverage for PLHIV, SW and MSM. We documented a methodology in this article to measure Commitment 6 and included SAS code for easy use with PHIA data sets containing HIV-related sub-population groups and social protection variables (Appendix 2).

Other nationally representative surveys measure access to social protection benefits. However, few also include HIV testing or questions relevant to identifying belonging to relevant sub-populations groups. The Multiple Indicator Cluster Survey (MICS) is one such survey. It has been periodically conducted in more than 100 low- and middle-income countries by UNICEF to assess children and women’s well-being. Like the PHIA, MICS are nationally representative surveys administered to individuals in households. The MICS 6 survey asks several questions about social protection, PLHIV, AGYW and OVC. The MICS 6 survey data sets have been released for Zimbabwe, Lesotho, the Democratic Republic of the Congo and Punjab province in Pakistan. The MICS survey does not ask questions that allow respondents to identify as MSM or SW [39]. Demographic and Health Surveys (DHS) are also nationally representative cross-sectional surveys that include HIV testing and identify the various population groups of interest. Although DHS surveys have been conducted in 90 countries, allowing for significant cross-country comparisons, they unfortunately do not capture information on social protection. Neither do they capture information on MSM [40].

Other sources explored that capture social protection coverage estimates in countries included the World Social Protection Database, hosted by the ILO. The database compiles and disseminates social security data by country and population group. It presents the proportion of the population “receiving at least one contributory or non-contributory cash benefit, or actively contributing to at least one social security scheme” among children, mothers with newborns, persons with severe disabilities, unemployed, older persons, vulnerable persons and the poor [41]. Another is the World Bank’s Atlas of Social Protection Indicators of Resilience and Equity, which compiles global social protection and labour indicators. None of the two capture HIV-related information [24].

There are several limitations to our study results. First, receipt of social protection benefits is self-reported, linked to a household and could not be verified independently. Respondents reporting that they or their households received benefits does not confirm that the respondent specifically received the benefit. However, it is assumed that household members shared the benefits a household received. Second, the data available to estimate the proportion of SW and MSM who reported receiving social protection benefits is small. As a result, the CI are wide, for SW and MSM, limiting the precision of our analyses. Third, the PHIA data sets that included HIV status and social protection information were only publicly available for Eswatini, Malawi, Tanzania and Zambia at the time of this analysis, limiting estimates outside these countries. Last, the PHIA data sets may not effectively capture receipt of social protection benefits for SW and MSM who have no fixed residence or did not identify as such or feel comfortable disclosing their social identity. People in prison, in the military, hospital, boarding schools and other institutions are not included in household-based surveys. However, our results are comparable with those obtained by the ILO [6]. We recommend that surveys being conducted among key populations include questions to capture social protection coverage.

## CONCLUSIONS

This study measured UNAIDS Fast-Track Commitment 6, “Ensuring that by 2020, 75% of people living with, at risk of or affected by HIV benefit from HIV-sensitive social protection.” In some of the highest HIV prevalence countries of the world (Eswatini, Malawi, Tanzania and Zambia), access to social protection benefits by the general population, PLHIV, AGYW, OVC, SW and MSM were lower than the global average of 45% and far short of 75% indicated in Commitment 6. More OVC, AGYW and PLHIV reported receiving social protection services greater than or equal to the general population; fewer SW and MSM did. Including SW, MSM and other key populations in population-based surveys that measure HIV and social protection is required to better estimate the prevalence of social protection benefits for these population groups. Data on access to social protection benefits by people living with, at risk of or affected by HIV are needed to better estimate their social protection coverage.

## Supporting information

Access to social protection Table 1

Access to social protection Table 2

Access to social protection Table 3

## Data Availability

We obtained the PHIA data sets from the PHIA Project website at https://phia-data.icap.columbia.edu/files.

## Declarations

We acknowledge technical and financial support from the Joint United Nations Programme on AIDS (UNAIDS). Olivia Keiser was supported by the Swiss National Science Foundation (grant no 163878). We also thank the participants UNAIDS convened in Geneva 16 - 17, December 2019, to review and input into the conceptual framework for developing a methodology to estimate social protection coverage among people living with, at risk of and affected by HIV.

## Ethics approval

The study did not require ethical clearance because the data are publicly available and deidentified.

## Consent to participate

Not applicable

Consent for publication (consent statement regarding publishing an individual’s data or image) Not applicable

Availability of data and material (data transparency)

PHIA Project website at https://phia-data.icap.columbia.edu/files

Code availability (software application or custom code)

Appendix 2. SAS Code.

## Authors’ contributions

David Chipanta and Audrey Pettifor conceived the study. Data preparation and analyses were performed by Jessica E Justman, Rachel M. Bray, Monique C. Millington, David Chipanta, Audrey Pettifor, Jesse Edwards, Danielle Giovenco, Hillary Mariko Topazian, Janne Estill and Olivia Keiser. The first draft of the manuscript was written by David Chipanta. All authors reviewed and commented on previous drafts. All authors have read and approved the final manuscript.

## Additional files

Appendix 1. Key variable names and questions used (PHIA survey instruments)

Information on file format. Brief description of file content.

