## Supplementary material for "Access to social protection by people living with, at risk of, or affected by HIV in Eswatini, Malawi, Tanzania and Zambia - results from Population-Based HIV Impact Assessments": Access to social protection Table 1

**Table 1. Per capita gross domestic product, Gini index, poverty ratio, HIV estimates and proportion of people who reported accessing any social protection benefit, by country**

| Indicator | Eswatini | Malawi | Tanzania | Zambia |
|---|---|---|---|---|
| Population[1] | 1 148 130 | 18 628 747 | 58 005 463 | 17 861 030 |
| Per capita gross domestic product (GDP) (current USD)[1] | 3894.68 | 411.55 | 1122.12 | 1305.06 |
| Gini index World Bank estimate[1] | 54.6 | 44.7 | 40.5 | 57.1 |
| Poverty head count ratio at national poverty lines (% of population)[1] | 58.9 | 51.5 | 26.4 | 54.4 |
| Number of adults and children living with HIV[2] | 200 000 [190 000– 220 000] | 1 100 000 [960 000–1 100 000] | 1 700 000 [1 500 000– 1 800 000] | 1 200 000 [1 200 000– 1 300 000] |
| HIV prevalence rate of adults ages 15 to 49 [2] | 27.0 [24.6–28.7] | 8.9 [7.6–9.6] | 4.8 [4.1–5.3] | 11.5 [10.9–12.1] |
| HIV prevalence among young women[2] | 12.3 [5.7–16.2] | 4.2 [2.2–5.8] | 2.2 [1.0–3.3] | 5.5 [2.8–7.4] |
| HIV incidence per 1000 population (adults 15–49 | 9.77 [7.79–12.44] | 3.71 [3.13–4.23] | 2.57 [2.27–2.87] | 6.03 [5.12–7.28] |
| Orphans due to AIDS ages 0 to 17[2] | 43 000 [36 000–49 000] | 460 000 [390 000–550 000] | 860 000 [740 000–990 000] | 350 000 [280 000–440 000] |
| HIV testing and treatment cascade | | | | |
| Percentage of PLHIV who know their status[2] | >95% [91–>95] | 90% [81–95] | 83 % [75–90] | 90% [85–>95] |
| Percentage of PLHIV who are on antiretroviral therapy[2] | >95% [88–>95] | 79% [71–84] | 75% [67–81] | 85% [80–92] |
| Percentage of PLHIV who have suppressed viral loads[2] | 92% [85–>95] | 72% [65–77] | 69% [62–74] | 77% [72–82] |
| Coverage of pregnant women who receive antiretrovirals for prevention of mother-to-child transmission (%)[2] | >95% [88–>95] | >95% [77–>95] | 92% [72–>95] | 86% [69–>95] |
| SW population size estimate (#)[2] | 4000 | 36 400 | 155 500 | 18 000 |
| MSM population size estimate (#)[2] | 2400 | 42 600 | 49 700 | 6500 |
| Social protection coverage in at least one area[3] | --- | 21.3 | --- | 15.3 |

[1] World Bank Population and GDP per capita (2019); GINI index for Eswatini (2016), Malawi (2015), Tanzania (2017) and Zambia (2015); poverty head count data for Eswatini (2015), Malawi (2016), Tanzania (2018) and Zambia (2015) (https://data.worldbank.org/indicator/SP.POP.TOTL?locations=SZ-MW-TZ-ZM).

[2] AIDSinfo [Internet]. Geneva: Joint United Nations Programme on HIV/AIDS.

(https://aidsinfo.unaids.org/).

[3] International Labour Organization (ILO). World social protection report 2017–2019: Universal social protection to achieve the Sustainable Development Goals. Geneva: ILO; 2017. Areas include child and family benefits, unemployment support, and health protection.

--- Data not available.
