## Supplementary material for "Access to social protection by people living with, at risk of, or affected by HIV in Eswatini, Malawi, Tanzania and Zambia - results from Population-Based HIV Impact Assessments": Access to social protection Table 2

**Table 2. Sample distribution by country and population group, unweighted percentage (%) and size, PHIA**

| Population group | Eswatini (2016–2017) | | Malawi (2015–2016) | | Tanzania (2016–2017) | | Zambia (2016) | |
|---|---|---|---|---|---|---|---|---|
| | **%** | **n** | **%** | **n** | **%** | **n** | **%** | **n** |
| Adults ages 15–59 years | | | | | | | | |
| PLHIV–female † | 20.1 | 1, 918 | 8.8 | 1,477 | 4.2 | 1192 | 8.8 | 1,688 |
| PLHIV–male † | 9.1 | 870 | 4.1 | 680 | 1.8 | 517 | 4.1 | 779 |
| MSM | --- | --- | 0.8 | 161 | 0.2 | 67 | 0.2 | 51 |
| SW–female | --- | --- | 1.0 | 200 | 2.7 | 803 | 0.4 | 76 |
| SW–male | --- | --- | ‡0.2 | 32 | 0.4 | 133 | ‡0.1 | 31 |
| AGYW | 19.7 | 2, 013 | 21.5 | 4,102 | 20.3 | 6,031 | 21.6 | 4,587 |
| Totals | 100.0 | 10,233 | 100.0 | 19,106 | 100.0 | 29,638 | 100.0 | 21,278 |
| Children ages 0–17 years | | | | | | | | |
| OVC | 27.8 | 2, 573 | 19.1 | 4,471 | 20.3 | 7,388 | 22.0 | 6, 094 |
| Total | 100.0 | 9, 271 | 100.0 | 23,432 | 100.0 | 36,376 | 100.0 | 27,655 |

† Denominators for PLHIV percentages were 9,556 in Eswatini, 16,698 in Malawi, 28,347 in Tanzania and 19,113 in Zambia, and excluded people who did not test for HIV or received indeterminate HIV test results.
--- Results had fewer than 25 adults identified during the survey and were suppressed.
‡ Estimate based on 25–49 persons/observations and should be interpreted with caution.
