## Supplementary material for "Access to social protection by people living with, at risk of, or affected by HIV in Eswatini, Malawi, Tanzania and Zambia - results from Population-Based HIV Impact Assessments": Access to social protection Table 3

**Table 3. Proportion reporting any external economic support, last 12 months by country and population group, PHIA (%, 95% CI, sample size)**

| Population | Eswatini (2016–2017) §[ş] | | | Malawi (2015–2016) § | | | Tanzania (2016–2017) § | | | Zambia (2016) § | | |
|---|---|---|---|---|---|---|---|---|---|---|---|---|
| Total population ages 15–59 | 39.6 | [36.8, 42.5] | 10,233 | 14.6 | [13.4, 15.9] | 19 106 | 8.8 | [7.9, 9.7] | 29 638 | 7.7 | [6.7, 8.8] | 21 278 |
| PLHIV - female | 39.4 | [35.7, 43.1] | 1,918 | 14.5 | [12.2, 16.8] | 1477 | 13.6 | [10.8, 16.5] | 1,192 | 7.3 | [5.7, 8.8] | 1,688 |
| PLHIV- male | 37.3 | [32.8, 41.8] | 870 | 14.8 | [11.5, 18.0] | 680 | 10.8 | [7.1, 14.4] | 517 | 6.4 | [4.5, 8.3] | 779 |
| MSM | -- | | | 11.8 | [6.5, 17.1] | 161 | 9.8 | [0.0, 20.0] | 67 | 7.0 | [0.4, 13.7] | 51 |
| SW - female | -- | | | 12.9 | [7.4, 18.4] | 200 | 11.7 | [8.5, 14.9] | 803 | 6.4 | [0.6, 12.3] | 76 |
| SW - male | -- | | | ‡4.2 | [0.0, 13.1] | 32 | 7.8 | [1.8, 13.8] | 133 | ‡11.9 | [0.0, 25.9] | 31 |
| AGYW | 44.8 | [41.0, 48.6] | 2,013 | 15.0 | [13.2, 16.7] | 4,102 | 8.7 | [7.5, 10.0] | 6,031 | 7.9 | [6.5, 9.3] | 4,587 |
| OVC ¶ | 44.8 | [41.5, 48.1] | 2,573 | 17.4 | [15.5, 19.3] | 4,471 | 6.1 | [4.9, 7.3] | 7,388 | 14.4 | [12.3, 16.4] | 6,094 |

§ Combined external economic support to the household in the last three or 12 months: social pension, material or financial support for shelter, food assistance provided at the household or external institution, income generation support in cash or kind (e.g. agricultural inputs), material support for education (e.g. uniforms, school books, education, tuition support, bursaries), assistance for school fees, cash transfer (e.g. pension, disability grants, child grant) or other. Denominator: all interviewed adults ≥ 15 years included in key population group definitions. Numerator: those who indicated social protection coverage.

¶ Combined school, social, material, emotional and medical support. Denominator: children < 18, conditional on whether the child, natural mother, and/or natural father has been very sick for at least three months during the past 12 months (too sick to work or do normal activities). Numerator: those who indicated receipt of child support in the last 12 months.

[ş]No 12-month variable included in the data set.

--- Results had fewer than 25 adults identified during the survey and were suppressed.

‡ Estimate based on 25–49 persons/observations and should be interpreted with caution.
